# Impacts of housing condition interventions on mental health: A systematic review

**DOI:** 10.1101/2025.04.04.25325141

**Authors:** Eleanor Shiels, Mathew Toll, Kaya Grocott, Kate Mason, Rebecca Bentley, Ang Li

## Abstract

Housing conditions have been increasingly recognised as a critical determinant of health, with emerging evidence highlighting their impact on mental well-being. However, there is a lack of research syntheses to provide a comprehensive overview of the mental health benefits of interventions aimed at improving indoor home environments. This review synthesises studies examining the effects of housing quality improvements on mental health, identifying which interventions, if any, are associated with improved mental well-being and to what extent. A comprehensive search of MEDLINE, Web of Science and PsycINFO was conducted for interventional studies published between January 2013 and April 2025. Results were analysed using frequency and thematic analyses, and the effect sizes of each intervention’s impact on mental health were summarised. The risk of bias was assessed using the ROBINS-I tool for non-randomised interventional studies. Fifteen studies met the inclusion criteria and were categorised according to the type of intervention. Findings showed significant mental health benefits following improvements to lighting, heating, kitchen, bathrooms, windows, and water heaters, with mixed evidence of mental health benefits from door installation, insulation, ventilation, and fabric works. The effectiveness of some housing modifications appears to be dependent upon parallel interventions. Most studies had a moderate to severe risk of bias. Further research should explore the generalisability of findings to broader populations, disentangle the health effects of individual interventions from confounders, and examine long-term mental health outcomes of housing condition interventions.

## Introduction

Mental distress and mental illness have been steadily rising for decades, with a noticeable acceleration since the onset of the COVID-19 pandemic (Li et al., 2025; Liao et al., 2021). Housing plays a central role in shaping people’s mental health and wellbeing, as evidenced by a growing body of observational studies that have established links between housing security, affordability, and quality, and the wellbeing of residents (Arundel et al., 2022; Jacobs et al., 2010; Li et al., 2022; Thomson et al., 2001; Thomson et al., 2013). Indoor temperature and other physical conditions of people’s homes have a known link to physical and mental health (Bentley et al., 2023; Thomson et al., 2013; World Health Organization, 2018). Living in a house of poor quality with inadequate ventilation or insulation can be hazardous for residents’ health and wellbeing (Gatto et al., 2024; Li et al., 2023; Vaid, 2023). These links between housing and health have contributed to a move by many governments towards legislating for minimum housing standards (e.g. New Zealand’s Healthy Homes Guarantee or the United States’ Department of Housing and Urban Development’s minimum property standards) and the implementation of initiatives and programs to improve the physical quality of housing and remediation of home environment hazards.

However, the evidence base remains limited on whether, and to what extent, improvements in housing conditions affect the health and wellbeing of resident (Thomson et al., 2013). Despite the increasing need for housing standards to promote population health, there has been a lack of comprehensive synthesis of evidence that examines how housing interventions - programs, initiatives, or modifications – impact on mental health and wellbeing as well as the magnitude of these effects. While there have been reviews on the association between health and housing quality conducted in the past (Thomson et al., 2001; Thomson et al., 2013), previous reviews date back a decade and all had inconclusive results, which highlights the need for an updated overview of the literature. There is also a paucity of reviews focused on mental health as the primary outcome and causal study designs.

This systematic review aims to synthesis the research evidence on the association between housing-focussed interventions and mental health from recent interventional studies. This review will enable a better understanding of how housing interventions can be effectively utilised to improve people’s mental health and prevent mental issues that may result from an unhealthy or unsafe living environment. This review will address the gap in the literature on housing research focusing on the link between mental health outcomes and housing interventions across domains, to inform future decisions by governments and policymakers.

## Methods

### Search strategy

MEDLINE, Web of Science, and PsycINFO were searched on 26 April 2025 for housing intervention studies published between 2013 and 2023, inclusive, and searches were restricted to human studies published in English. Search terms consisted of four blocks of search terms: dwelling (“house” or “housing” or “dwelling” or “home” or “residential”); changes in housing condition (“decrease” or “increase” or “evaluation” or “change” or “intervention” or “improve” or “effect” or “impact”); mental health (“mental” NEAR/2 (“health” OR “illness”) or “depression” or “anxiety” or “psychological distress”); and indoor environment (“indoor” or “thermal” or “heat” or “insulation” or “mould” or “double glazing” or “pest” or “damp” or “lightning” or “retrofit” or “ventilation” or “refurbish” or “overcrowd” or “crowding” or “ noise” or “asbestos” or “lead” or “radon” or “sanitation” or “safety” or “injury” or draught” or “allergen” or “renovation” or “repair”). We used the Boolean operators ‘AND’ and ‘OR’ as well as wildcards to allow for variations of the search words and included alternative spellings and synonyms. The review was registered on Prospero (ID CRD42023445233).

### Inclusion and exclusion criteria

Studies were excluded if they did not include one of the four components. Qualitative studies and studies that did not include an intervention were excluded. Studies examining interventions to address adverse effects of lead, poor air quality, or radon were not included because these are environmental hazards, rather than poor quality housing. Housing settings that were not individual residences, such as residential aged care facilities were excluded. Studies with an exclusively child (under 18 years) population were excluded, and where studies reported outcomes for both children and adult participants separately, only the results for adults were used. Covidence was used for both abstract and full text screening.

### Study selection

Studies that used experimental or quasi-experimental approaches to examine the mental health effects of housing improvements were included. Outcome measures included any quantitative measure of mental health, including wellbeing, quality of life, depression, stress, and anxiety. The studies were required to provide a measurement of mental health either pre- and post-intervention for the intervention group, or post-intervention for both intervention and control/comparison groups. Housing interventions were defined as a change or renovation in a residence, which was implemented by a study or program, such as increased lighting, central heating installation, new doors and windows, kitchen and bathroom renovations, major repairs and upgrades and asbestos removal. Housing interventions did not include changes to workplaces or other non-residential spaces, or changes to the surrounding environment such as tree planting.

### Data extraction

A data extraction sheet was created with the following data extracted from each study: Citation (Journal, authors, title, year of publication, volume, page numbers/article number); Methods (Aim, study design, setting, study period, intervention(s), type of measurement used; any variables controlled for during analysis); Demographics (Age, gender, household size, type of housing); Sample size (before and after follow-up); Results (Overall mental health, anxiety, depression, stress) for each of the following: Control group before/after, intervention group before/after, mean difference for both control and intervention groups, significance of the mental health change in each group. Data was extracted by two reviewers independently, and disagreements were resolved with the team. When a specific piece of information was unavailable, but similar data was available then that data was extracted instead. If a specific piece of information was unavailable, and no similar data was available then these fields were left blank. Results of the process were reported in the study summary and synthesis.

### Study quality assessment

The ROBINS-I tool for non-randomised interventional studies was used to conduct a risk-of-bias assessment for all included studies (Jonathan et al., 2016). A scale with values of low, moderate, serious, and critical was used to assess each domain and the overall risk of bias.

### Methods of synthesis

Where possible, studies were categorised based on the intervention target (e.g. lighting, heating). The data were synthesised narratively using frequency and thematic analyses, with substantive findings analysed and synthesised separately for each intervention category. Given the high heterogeneity in study design, interventions, and measurements, meta-analysis was not appropriate for this review.

Due to heterogeneity in measurements between studies, only the primary or “overall” mental health score was extracted unless otherwise specified. Where multiple outcomes are listed, the study did not have a main mental health measure and instead had several measures of different aspects of mental health. The mean difference between pre- and post-intervention mental health scores and significance level were included in the results. For studies that did not report this value, the mean difference was calculated by taking the mean post-intervention mental health and subtracting the mean pre-intervention mental health.

Some studies had a control or comparison group while others only had an intervention group; where applicable, results are listed for both groups.

### Categorisation of interventions

Interventions were grouped into heating (including central heating), insulation (including cavity wall insulation, external wall insulation, loft/attic insulation, roof insulation, exterior duct insulation and air sealing), ventilation (including kitchen and bathroom exhaust vented externally, new building ventilation on roof, mechanical ventilation), lighting (including increasing light levels), fabric works (including stabilizing and restoring concrete structure, painting, repairs to walls), windows (including replacement with new double glazed windows), water heating (including replacement with new water heater), kitchen and bathroom renovation (including installation of new kitchen and bathroom), and doors (including new door installation), based on the definitions provided by the studies.

## Results

### Reviewed studies

Initial searches identified 772 studies after deduplication. After title and abstract screening, 20 papers remained. Six of these were excluded at the full text screening stage; four were qualitative studies, one did not have an intervention and was purely observational, and one did not report the relationship between mental health and the housing interventions. Detailed reasons for exclusion of each study are reported in the Appendix (Table A1). The PRISMA flow diagram is displayed in Figure 1, which was generated using Covidence. A total of fifteen papers were included in the review after screening.

**Figure 1.**
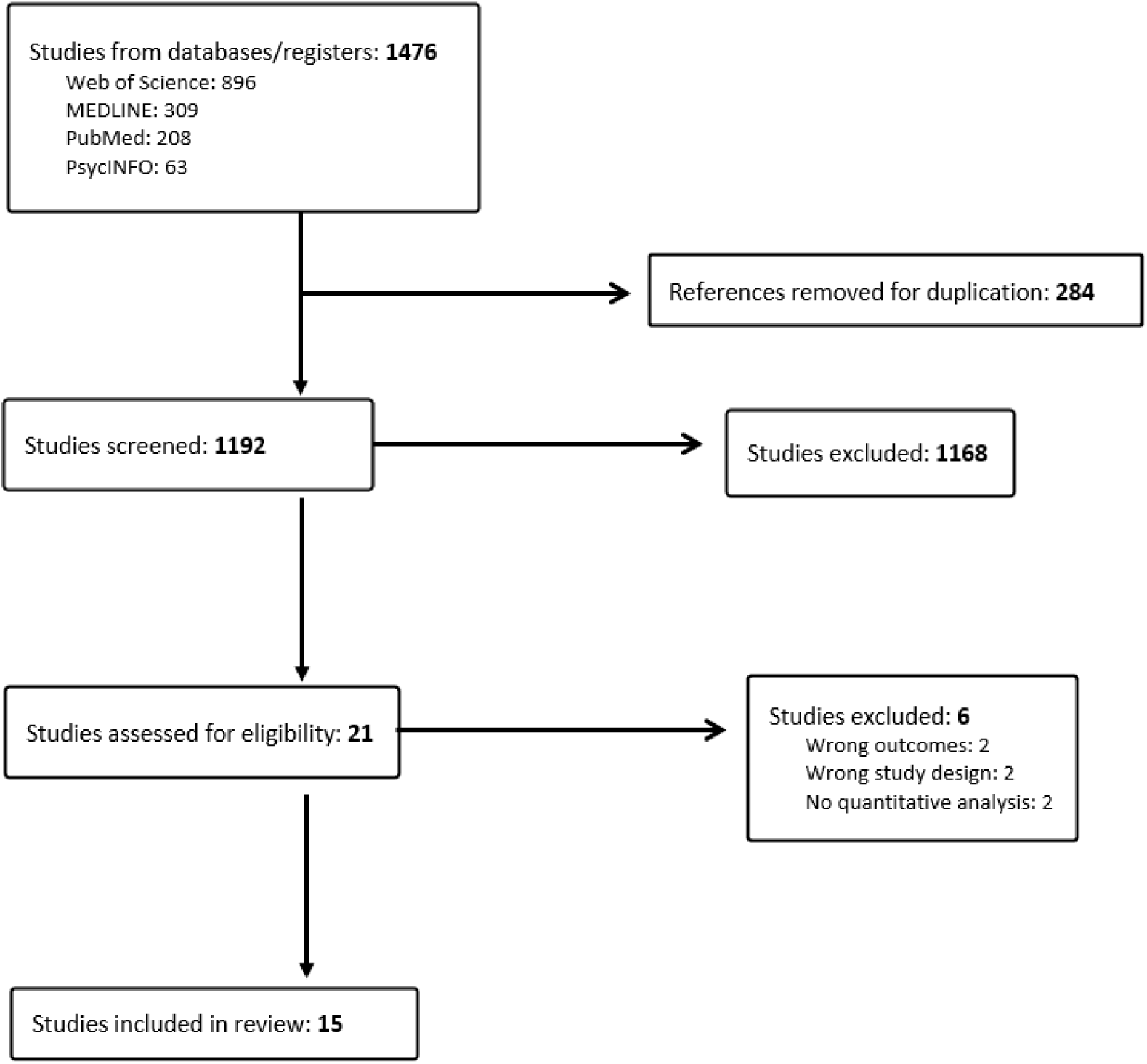
PRISMA flow diagram.

### Study designs

Two crossover studies, seven cohort studies, and six cross-sectional studies were eligible for inclusion.

### Demographics

Table 1 presents the demographics of participants in each included study. All but three studies reported either an age range or mean age in their sample, and all but one study reported the gender ratio. Overall, most studies reporting a mean age had a population that was primarily over 50 years old and female. This may be due to societal factors resulting in women having less financial security in their lifetimes, and therefore being more likely to live in lower quality housing, particularly in old age. Older people may also be more likely to live in low quality housing requiring renovations due to retirement or decreased ability to work. Six studies assessed the effects of interventions on public housing.

**Table 1.** Demographics of study participants.

| Author Year, Country | Group (control or intervention) | Age group (range or mean) | Gender ratio (% female) | Initial sample size | Final sample size | Type of housing |
| --- | --- | --- | --- | --- | --- | --- |
| *Tonn 2021, USA (Tonn et al., 2021)* | Control | 59 years | 79% | 152 | 100 | Owner-occupied homes |
|  | Intervention | 56 years | 77% | 99 | 79 |  |
| *Falkenberg 2019, Norway (Falkenberg et al., 2019)* | Control | 77 years | 47% | 30 | 29 | Private homes |
|  | Intervention | 77 years | 57% | 30 | 29 |  |
| *Tonn 2023, USA (Tonn et al., 2023)* | Control | 60 years | 60% | 1273 | 700 | Multifamily units without weatherisation |
|  | Intervention | 60 years | 73% | 742 | 309 | Multifamily units that received weatherisation in the past 5 years |
| *Nagare 2021, USA (Nagare et al., 2021)* | Intervention only | 21-77 years | 55% | 20 | 20 | Luxury apartment complex |
| *Jacobs 2014, USA (Jacobs et al., 2014)* | Intervention only | 38 years | 93% | 57 | 27 | Low-income housing |
| *Jacobs 2015, USA (Jacobs et al., 2015)* | Control | 33 years | 90.1% | 91 | Not stated | Public or federally subsidised housing |
|  | Intervention | 43 years | 79.5% | 298 | Not stated |  |
| *Breyse 2015, USA (Breyse et al., 2015)* | Control | 33-86 years | 70% | 40 | 40 | Low-income |
|  | Intervention | 33-86 years | 70% | 40 | 40 | Public housing |
| *Curl 2015, UK (Curl & Kearns, 2015)* | Control | Not stated | Not stated | 602 | 602 | Public housing |
|  | Intervention | Not stated | Not stated | 1933 | 1334 |  |
| *Grey 2017, UK (Grey et al., 2017)* | Control | Not stated | 59.7% | 852 | 418 | Private homes |
|  | Intervention | Not stated | 58.8% | 656 | 364 |  |
| *Poortinga 2017, UK (Poortinga et al., 2017)* | Intervention only | 18+ years | 69% | 2075 | 1709 | Public housing |
| *Sawyer 2022, UK (Sawyer et al., 2022)* | Intervention only | 22-93 years | 71.4% | 149 | 78 | Private homes |
| *Curl 2015, UK (Curl et al., 2015)* | Control | Not stated | 62.8% | 602 | 602 | Public housing |
|  | Intervention | Not stated | 60.5% | 1933 | 1334 |  |
| *Bray 2017, UK (Bray et al., 2017)* | Intervention only | 25+ years | 75.6% | 389 | 228 | Public housing |
| **Shishegar 2022, USA (Shishegar & Boubekri, 2022)** | Intervention only | 76.81 years | 76% | 21 | 21 | Independent living communities |
| **Page 2025 \*citation\*** | Control | 74.85 years | 67% | 493 households | 649 individuals (ITT analysis) | Low-income private homes |
|  | Intervention | 74.82 years | 63% | 491 households | 664 individuals (ITT analysis) | Low-income private homes |

### Mental health outcome measures

Outcome measures included mental wellbeing, quality of life, depression, stress, and anxiety (Table 2). These are measured using a standard mental health related questionnaire such as 36-Item Short Form Survey (SF-36), Short form 12 Item Health Survey (SF-12), Patient-Reported Outcomes Measurement Information System (PROMIS), Veterans RAND 12-Item Health Survey (VR-12), Short Warwick-Edinburgh Wellbeing Scale, and Geriatric Depression Scale (GDS) or measures designed specific for the study such as the number of days in the past 30 days when mental health was not good and scores of six indicators of poor mental health including “Worthless”, “Everything was effort”, “Hopeless”, “Restless”, “Nervous”, “Sad”. For most studies, an overall mental health measurement was recorded, which was the primary mental health outcome used in the study. Some studies did not record an overall mental health measurement but reported a score in one or more other mental health categories, while other studies stratified results by intervention. Details of the outcome measures and measurements used were extremely heterogeneous, and few studies used the same measure as any other. Many studies used a reverse scale, in which a higher score indicates worse mental health.

**Table 2.**
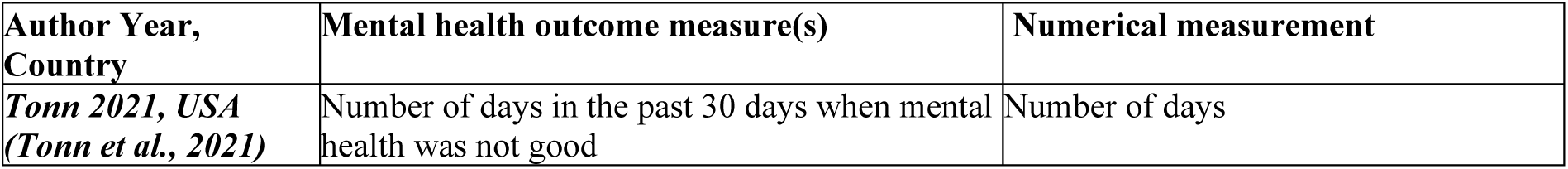
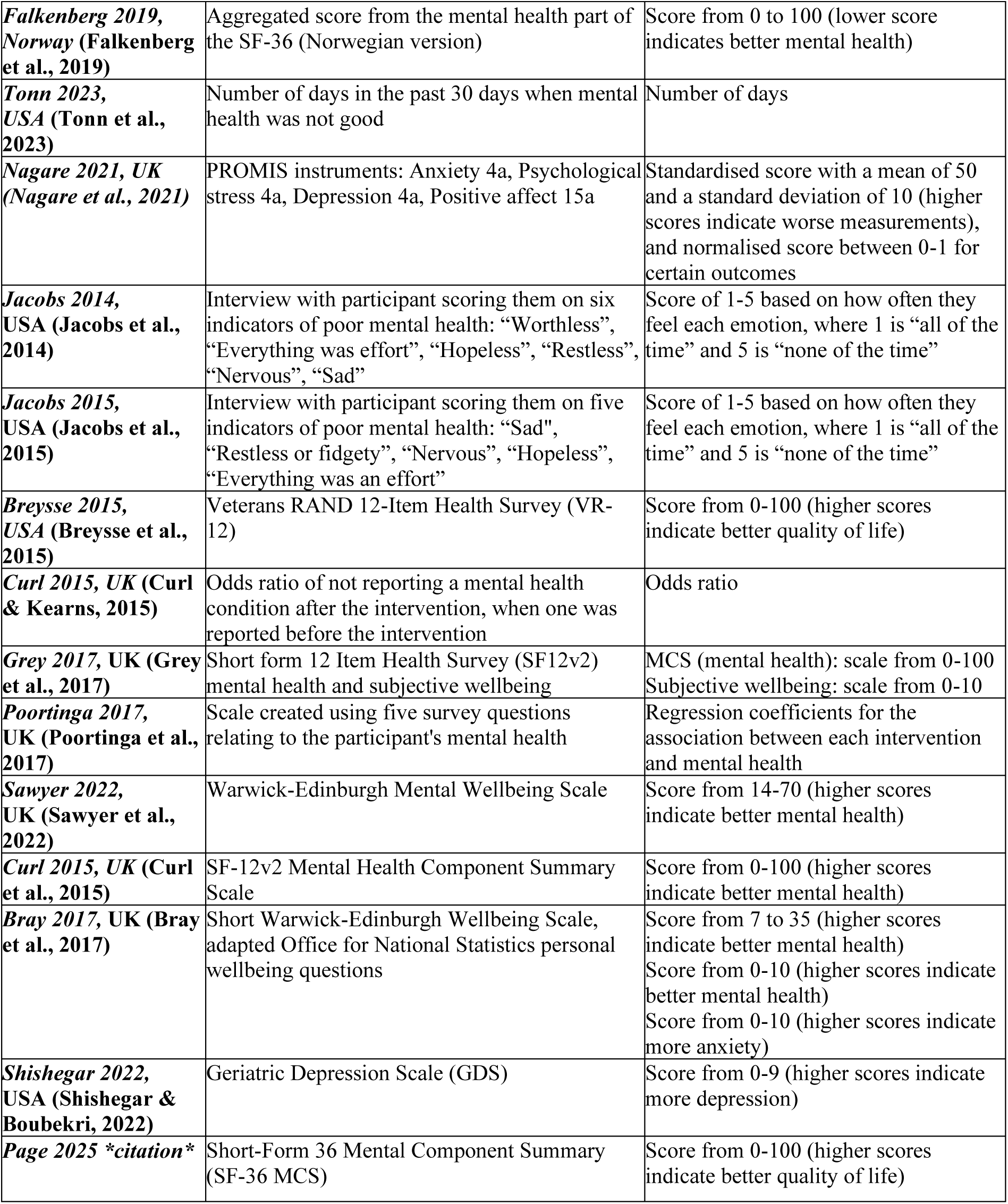
Outcome measures.

### Housing interventions

The most common interventions reported were heating (9), insulation (7), window replacement (6), ventilation (4), water heating (4), new doors (3), kitchen and bathroom renovations (3), and lighting (3). Interventions that were present in fewer than three studies were included in the table of results but not the narrative synthesis.

### Effects of interventions

Table 3 reports the study outcomes and findings. The results based on the main measure of mental health given in each study are reported. Some studies reported more than one main measure of mental health, and where applicable, these have been listed separately. The effect of interventions of mental health and related measures are synthesised on the basis of studies that examined interventions on one dimension of housing and those that examined interventions on a cluster of conditions. Three studies evaluated a single treatment and all of these evaluated lighting-based interventions; two interventions provided electrical lighting infrastructure and programmed their daily lighting schedule for older persons and found improvement for quality of life and mood measures (Falkenberg et al., 2019; Shishegar & Boubekri, 2022), and one found a positive mental health effect following the introduction of electrochromic glass windows (Nagare et al., 2021). The remainder of studies evaluated a combination of multiple dimensions of intervention on housing conditions. Seven (Breysse et al., 2015; Curl & Kearns, 2015; Curl et al., 2015; Jacobs et al., 2015; Poortinga et al., 2017; Sawyer et al., 2022; Tonn et al., 2021) of the twelve multiple intervention studies found positive effects on mental health from the improvements, while five studies (Bray et al., 2017; Grey et al., 2017; Jacobs et al., 2014; Page et al., 2025; Tonn et al., 2023) found insufficient evidence of an impact on mental health or had a different primary measure. The preponderance of the evidence from these combination studies indicated that insulation and heating interventions have a positive mental health effect (Breysse et al., 2015; Curl & Kearns, 2015; Jacobs et al., 2015; Poortinga et al., 2017; Sawyer et al., 2022; Tonn et al., 2021).

**Table 3.**
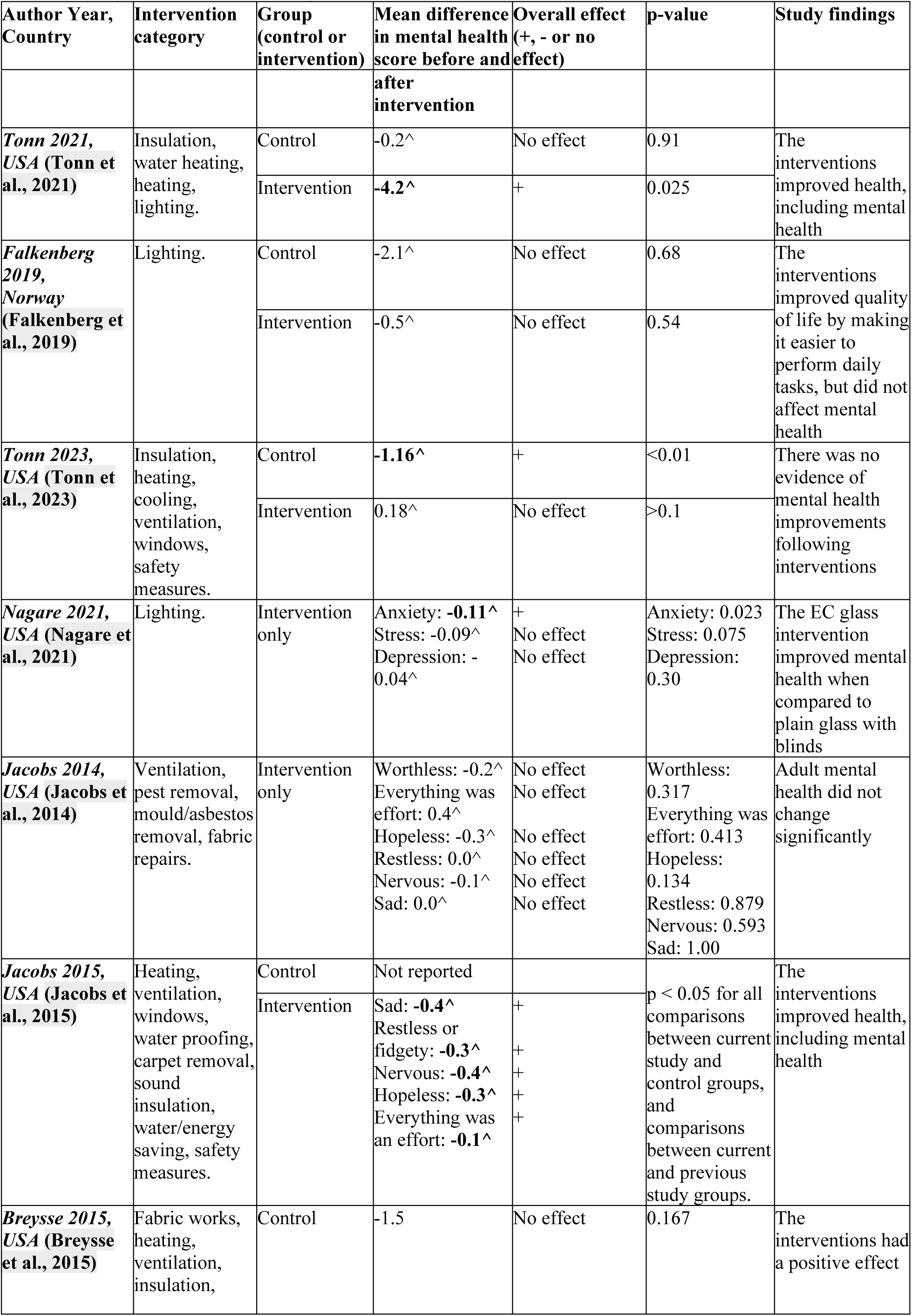
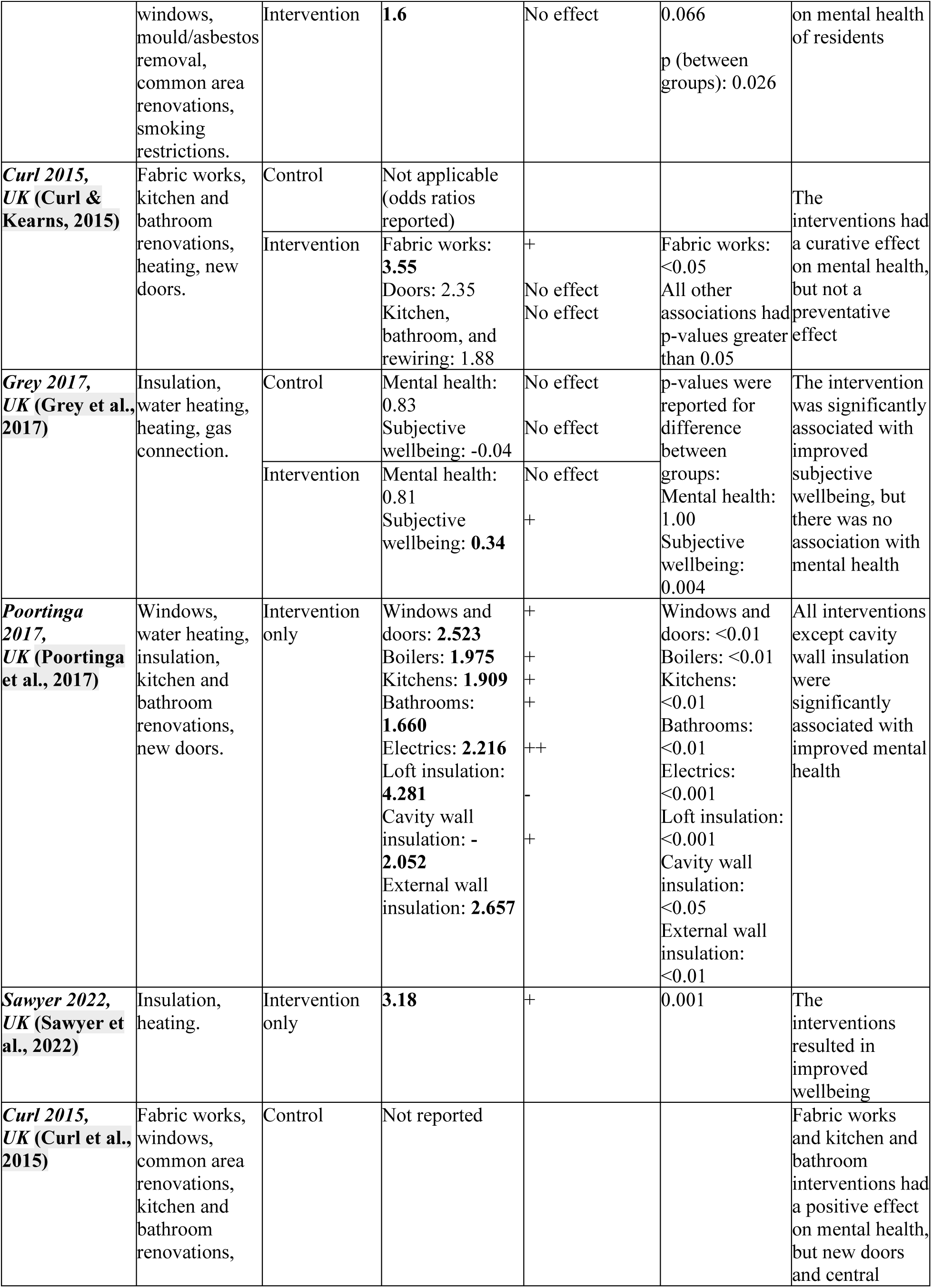
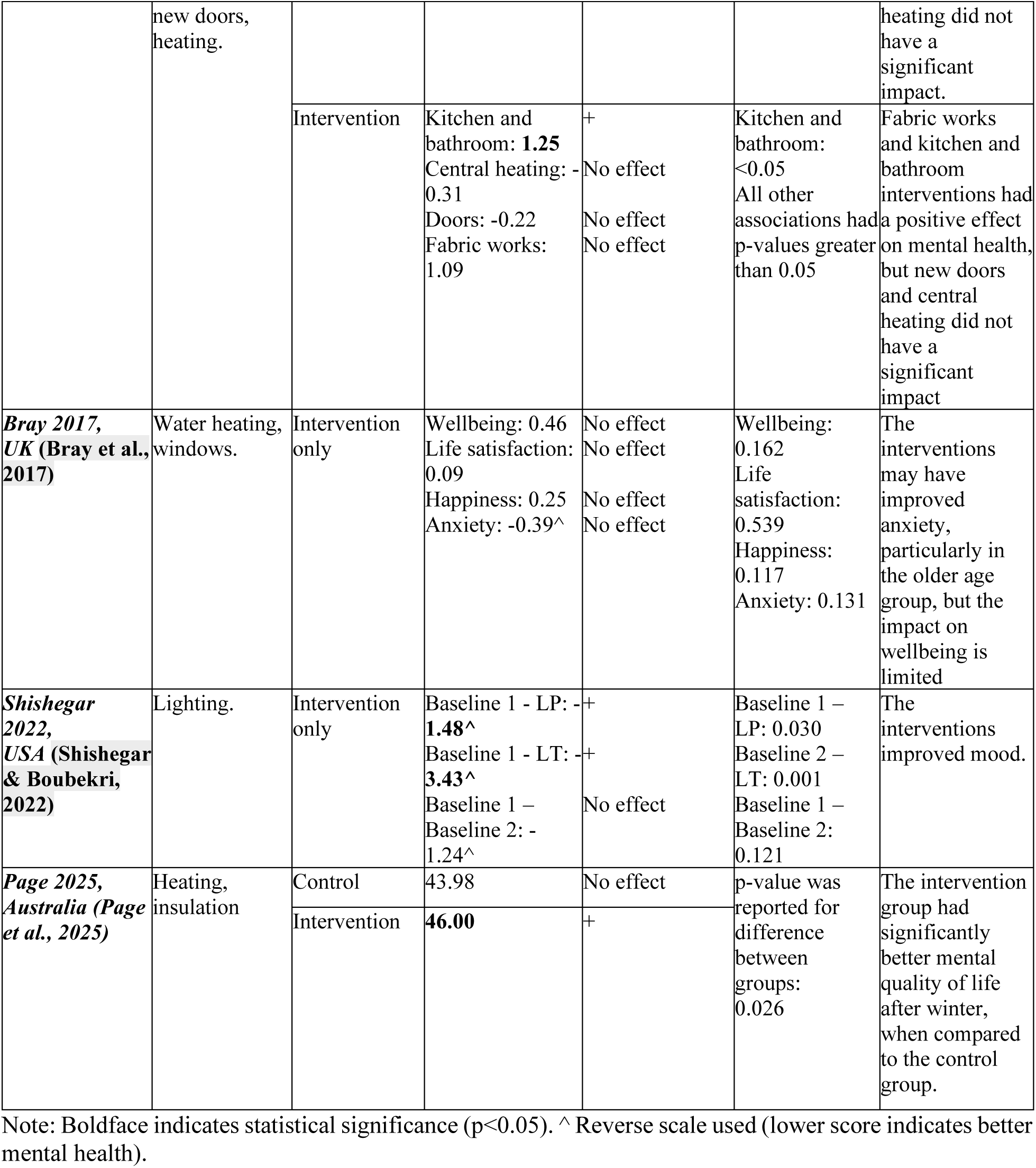
Study outcomes and findings.

Four studies (Breysse et al., 2015; Jacobs et al., 2015; Jacobs et al., 2014; Tonn et al., 2023) examined the mental health effects of ventilation, but only two of these (Breysse et al., 2015; Jacobs et al., 2015) studies found an improvement following the interventions. All four studies included other interventions, such as heating, mould removal, window replacement and insulation, suggesting that ventilation may not be impacting mental health alone, but may work in conjunction with other interventions to improve the overall home environment. This is supported by the findings of one study (Poortinga et al., 2017), which found that cavity wall insulation without ventilation appeared to worsen mental health, while external wall insulation with ventilation resulted in improvements in mental health.

The three studies (Breysse et al., 2015; Curl & Kearns, 2015; Curl et al., 2015) that reported fabric works as part of their interventions found fabric works was associated with improved mental health at follow-up, and one of these associations (Curl & Kearns, 2015) was statistically significant. The study that included fabric repairs (Jacobs et al., 2014) reported no significant change in mental health. The discrepancy in the mental health effects of fabric works may be because aesthetic interventions such as painting were more likely to result in mental health improvements than structural interventions such as repair of holes in walls, or that minor repairs were inadequate to substantially improve participants’ mental health and wellbeing.

Window replacement was a very common intervention, with six studies (Bray et al., 2017; Breysse et al., 2015; Curl et al., 2015; Jacobs et al., 2015; Poortinga et al., 2017; Tonn et al., 2023) including window replacement with energy efficient double-glazed windows, of which four studies (Breysse et al., 2015; Curl et al., 2015; Jacobs et al., 2015; Poortinga et al., 2017) found that interventions including window replacement resulted in improved mental health or wellbeing.

Four studies (Bray et al., 2017; Grey et al., 2017; Poortinga et al., 2017; Tonn et al., 2021) completed water heating upgrades, including the replacement of water heaters with new, more efficient ones. Three of these reported improvements in at least one measure of mental health, while one study (Bray et al., 2017) reported no significant changes.

Three studies included kitchen and bathroom renovations (Curl & Kearns, 2015; Curl et al., 2015; Poortinga et al., 2017), all of which found positive associations with mental health, and two (Curl et al., 2015; Poortinga et al., 2017) of these results were statistically significant. It is likely that improvements in mental health were at least partly due to improved internal appearance and functionality of the homes.

Three studies included new doors in their interventions, and all three stratified their results by intervention. Two of these studies (Curl & Kearns, 2015; Poortinga et al., 2017) found that new doors had a positive effect on mental health, while the other study found no effect.

### Risk of bias assessment

The ROBINS-I tool was used to assess the risk of bias for each study included in the review across domains of confounding, selection of participants, classification of interventions, deviations from intended interventions, missing data, measurement of outcomes, and selection of reporting (Jonathan et al., 2016), on a scale from low to critical to indicate the risk of bias present in the study (Table 4).

**Table 4.** Risk of Bias assessment.

| Author, publication year | Confounding | The selection of participants | Classification of interventions | Deviations from intended interventions | Missing data | Measurement of outcomes | Selective outcome reporting | Overall risk |
| --- | --- | --- | --- | --- | --- | --- | --- | --- |
| Tonn 2021 | Serious | Low | Low | Low | Low | Moderate | Moderate | Serious |
| Falkenberg 2019 | Low | Low | Low | Low | Low | Moderate | Moderate | Moderate |
| Tonn 2023 | Serious | Serious | Moderate | Low | NI | Serious | Moderate | Serious |
| Nagare 2021 | Low | Low | Low | Low | Low | Low | Low | Low |
| Jacobs 2014 | Moderate | Low | Low | Low | Low | Serious | Moderate | Serious |
| Jacobs 2015 | Moderate | Low | Low | Low | Low | Serious | Moderate | Serious |
| Breysse 2015 | Moderate | Serious | Low | Low | Low | Moderate | Moderate | Serious |
| Curl 2015 | Serious | Low | Low | Low | Moderate | Low | Moderate | Serious |
| Grey 2017 | Moderate | Low | Low | Moderate | Moderate | Moderate | Moderate | Moderate |
| Poortinga 2017 | Moderate | Low | Low | Low | Low | Moderate | Moderate | Moderate |
| Sawyer 2022 | NI | Low | Low | Low | Moderate | Serious | Moderate | Serious |
| Curl 2015 | Moderate | Low | Low | Low | NI | Low | Moderate | Moderate |
| Bray 2017 | Moderate | Low | Low | Low | Serious | Moderate | Moderate | Serious |
| Shishegar 2022 | Low | Low | Low | Low | Low | Low | Moderate | Moderate |
| Page 2025 | Moderate | Low | Moderate | Moderate | Moderate | Moderate | Moderate | Moderate |

Notes: NI (no information) was used if not enough information was available to make an assessment.

Eight of the fifteen studies (Bray et al., 2017; Breysse et al., 2015; Curl & Kearns, 2015; Jacobs et al., 2015; Jacobs et al., 2014; Sawyer et al., 2022; Tonn et al., 2023; Tonn et al., 2021) had a serious risk of bias in at least one domain, of which three studies has issues with confounding risks(Curl et al., 2015; Tonn et al., 2023; Tonn et al., 2021), and four studies had issues with measurement of outcomes, primarily due to the nature of the intervention or control group status being known when participants’ mental health is evaluated(Jacobs et al., 2015; Jacobs et al., 2014; Sawyer et al., 2022; Tonn et al., 2023). Two studies had serious risk of bias in the selection of participants due to enrolling participants who had already received intervention (Breysse et al., 2015; Tonn et al., 2023). One study had a serious risk of bias for missing data(Bray et al., 2017). Six studies (Curl et al., 2015; Falkenberg et al., 2019; Grey et al., 2017; Page et al., 2025; Poortinga et al., 2017; Shishegar & Boubekri, 2022) had a moderate risk of bias in at least one domain, but no domain had a serious risk. One study had an overall low risk of bias and can be considered comparable to a well-performed randomised trial (Nagare et al., 2021). Six of these studies used a baseline measurement of the intervention group but not the control/comparison group, and four studies had fewer than 50 participants in each group. Two studies did not report the period of data collection or analysis.

Of the lighting studies, one of them had a low risk and the other two had a moderate risk. Of the non-lighting studies, eight of twelve had a serious risk, four had a moderate risk of bias and none had a low risk. This difference is most likely because the lighting interventions were easier to reverse, faster to implement, and the groups could be randomised beforehand, while the non-lighting interventions were usually part of a program outside the study and the researchers were aiming to measure the effects of this.

## Discussion

The review found that most studies showed a significant and positive relationship between housing improvements and mental health. There was strong evidence supporting improvements to lighting, heating, kitchen, bathrooms, windows, and water heaters. Evidence supporting other interventions was inconsistent. Nine of the fifteen studies (Breysse et al., 2015; Curl & Kearns, 2015; Curl et al., 2015; Jacobs et al., 2015; Nagare et al., 2021; Poortinga et al., 2017; Shishegar & Boubekri, 2022; Tonn et al., 2021) found significant evidence supporting a link between housing interventions (including heating, lighting, windows, water heating, and kitchen and bathroom renovations) and mental health benefits. Three additional studies (Falkenberg et al., 2019; Grey et al., 2017; Sawyer et al., 2022) found improvements in other outcomes (e.g., quality of life and subjective wellbeing), but unchanged mental health.

Of the interventions on housing conditions, heating was the most common intervention in the included studies (Breysse et al., 2015; Curl & Kearns, 2015; Curl et al., 2015; Grey et al., 2017; Jacobs et al., 2015; Page et al., 2025; Sawyer et al., 2022; Tonn et al., 2023; Tonn et al., 2021). Four of these studies showed statistically significant improvements in mental health after heating improvements were completed, and three studies showed improvements in wellbeing (Howden-Chapman et al., 2023; Mishra et al., 2023; Page et al., 2025). Insulation was another common intervention, with seven studies (Breysse et al., 2015; Grey et al., 2017; Page et al., 2025; Poortinga et al., 2017; Sawyer et al., 2022; Tonn et al., 2023; Tonn et al., 2021) involving insulation improvements. Three studies (Breysse et al., 2015; Poortinga et al., 2017; Tonn et al., 2021) found that at least one type of insulation installed resulted in mental health improvements, and two additional studies (Grey et al., 2017; Sawyer et al., 2022) found improvements in wellbeing. Installation of double-glazed windows was found to have a positive mental health effect in four studies (Breysse et al., 2015; Curl et al., 2015; Jacobs et al., 2015; Poortinga et al., 2017). Improvements in ventilation were not implemented independently, and their effectiveness appears to be dependent upon parallel interventions (Breysse et al., 2015; Jacobs et al., 2015; Jacobs et al., 2014; Tonn et al., 2023). It is unclear whether fabric works or repairs affect mental health, and this is complicated by the heterogeneity of interventions that fall under these broad terms. The majority of the studies found water heating (three of four studies) ^14,22,23^, kitchen and bathroom renovations (two of three studies) (Curl et al., 2015; Poortinga et al., 2017), and new doors (two of three studies) (Curl & Kearns, 2015; Poortinga et al., 2017) were associated with increases in mental health, likely due to the direct improvement in safety and functionality of the home environment. Potential mechanisms for the translation of housing quality improvements to mental health gains suggested in the research literature can be analytically divided into somatic and psychological pathways: physical health gains from thermal comfort and reduced sleep disturbance, for instance, can lead to mental health co-benefits and direct psychological mechanisms include reduced stressors, increased sense of control, and dignity post-intervention.

These review findings are consistent with other studies that link indoor thermal conditions and energy efficiency to health (Bentley et al., 2023; Howden-Chapman et al., 2023; Mishra et al., 2023; Thomson et al., 2001), while also underscoring that heating, insulation, and window interventions can produce mental health gains. Complementing early systematic reviews on housing refurbishment and health outcomes (Thomson et al., 2001; Thomson et al., 2009), this review covers broader dimensions of housing condition interventions and provides effect size estimates to enable the comparison across interventions and consideration of clinical significance of mental health effects. The heterogeneity of the findings is related to the scope of the intervention and the targeted population.

The risk of bias assessment suggests that studies were largely at the moderate to serious risk of bias in the study design or conduct of the study. Three studies (Bray et al., 2017; Jacobs et al., 2014; Tonn et al., 2023) found that the interventions did not have a significant impact on any measure of mental health. The quality of evidence supporting a link between positive outcomes and lighting interventions is strong, with all lighting studies having low or moderate risk of bias. Studies of other intervention types are affected by moderate or serious risks of bias but no study in the corpus had a critical risk of bias that would exclude it from consideration for synthesis of evidence.

By synthesising interventional studies only, we reduce the influence of unmeasured confounding on our overall conclusions. While many studies were available to include in the study, few had a quasi-experimental design, meaning that they may have an increased risk of bias as participants may choose their interventions or there may not be any baseline measurement. Excluding these studies at the screening stage resulted in a stronger evidence base from which to draw conclusions.

This review has some limitations. First, the outcome measures were highly heterogeneous, and the quality of studies varied considerably. Most studies examined the effects of multiple interventions at once, which made it difficult to separate the effects of each intervention and draw conclusions about the magnitude and direction of the effect of specific interventions. Second, grey literature and studies in languages other than English were not included. Third, a meta-analysis was unable to be performed due to the heterogeneity of the data.

There was strong evidence supporting improvements to lighting, heating, kitchen, bathrooms, windows, and water heaters. Evidence supporting other interventions was inconsistent, and more research is required to determine if and under what circumstances these improvements are beneficial. As the populations in some studies were limited to older adults, further research is needed to determine whether these findings can be applied to the general population. Further research is required to understand the way different types of insulation affect mental health when installed with and without ventilation. As fabric works is a very broad category, these improvements could be broken down into smaller subcategories such as painting, over-cladding, etc. Further research should also be done on the effects of new doors on mental health over time, as initial improvements appear to wear off over time which may be due to perceived safety effects.

There is a long standing concern with housing affordability and security in many high-income countries contexts, though there have been recent calls to consider housing suitability for health promotion as a problem for housing policy (Dinmore et al., 2024). There is evidence supporting the mental health benefits of improvements to lighting, heating, kitchens and bathrooms, windows, and water heaters, which should be considered in housing and rental standards to support the health and wellbeing of residents. The importance of housing suitability was drawn into sharp relief with the recent COVID-19 pandemic that confined people within health harming housing conditions (Li et al., 2025) and with the anticipation of climate change (Li et al., 2024). These studies provide an evidence basis for the effect of housing condition interventions and support the case for policies requiring health promoting building codes and minimum standards for new and existing homes.

## Conclusion

This review summarises literature on the relationship between mental health and housing quality using intervention studies and compares the effect sizes of different housing condition interventions. With few reviews examining mental health as a primary outcome or synthesising evidence from studies using causal study designs, this study contributes to the understanding of mental health impacts of interventions, initiatives, and modifications of housing conditions. Mechanisms within the literature suggest indirect pathways via physical health and direct psychological processes that result in mental health gains. Consistent findings on the mental health benefits of improvements were found by studies examining lighting, heating, kitchens, bathrooms, windows, and water heaters, suggesting a stronger evidence basis for these types of housing interventions to be incorporated in programs and policies aimed at the prevention of mental disorders and the promotion of mental health. This review also underscores the importance of health-in-all policies that incorporate health protection and promotion in policy development outside of the silo of health policy.

## Data Availability

All data produced in the present work are contained in the manuscript.

## Registration

This review was registered on PROSPERO under ID CRD42023445233.

## Ethical considerations

There are no human participants in this article and informed consent is not required.

## References

Arundel, R., Li, A., Baker, E., & Bentley, R. (2022). Housing unaffordability and mental health: dynamics across age and tenure. International Journal of Housing Policy, 1–31. 10.1080/19491247.2022.2106541

Bentley, R., Daniel, L., Li, Y., Baker, E., & Li, A. (2023). The effect of energy poverty on mental health, cardiovascular disease and respiratory health: a longitudinal analysis. The Lancet Regional Health – Western Pacific, 35. 10.1016/j.lanwpc.2023.100734

Bray, N., Burns, P., Jones, A., Winrow, E., & Edwards, R. T. (2017). Costs and outcomes of improving population health through better social housing: a cohort study and economic analysis. Int J Public Health, 62(9), 1039–1050. 10.1007/s00038-017-0989-y

Breysse, J., Dixon, S. L., Jacobs, D. E., Lopez, J., & Weber, W. (2015). Self-reported health outcomes associated with green-renovated public housing among primarily elderly residents. J Public Health Manag Pract, 21(4), 355–367. 10.1097/phh.0000000000000199

Curl, A., & Kearns, A. (2015). Can housing improvements cure or prevent the onset of health conditions over time in deprived areas? BMC Public Health, 15(1), 1191. 10.1186/s12889-015-2524-5

Curl, A., Kearns, A., Mason, P., Egan, M., Tannahill, C., & Ellaway, A. (2015). Physical and mental health outcomes following housing improvements: evidence from the GoWell study. J Epidemiol Community Health, 69(1), 12–19. 10.1136/jech-2014-204064

Dinmore, H., Beer, A., Baker, E., & Bentley, R. (2024). Advancing a healthy housing policy agenda: how do policy makers problematise housing-related health issues? Journal of Social Policy, 1–18.

Falkenberg, H. K., Kvikstad, T. M., & Eilertsen, G. (2019). Improved indoor lighting improved healthy aging at home - an intervention study in 77-year-old Norwegians. J Multidiscip Healthc, 12, 315–324. 10.2147/jmdh.S198763

Gatto, M. R., Mansour, A., Li, A., & Bentley, R. (2024). A State-of-the-Science Review of the Effect of Damp-and Mold-Affected Housing on Mental Health. Environmental Health Perspectives, 132(8), 086001.

Grey, C. N., Jiang, S., Nascimento, C., Rodgers, S. E., Johnson, R., Lyons, R. A., & Poortinga, W. (2017). The short-term health and psychosocial impacts of domestic energy efficiency investments in low-income areas: a controlled before and after study. BMC Public Health, 17(1), 140. 10.1186/s12889-017-4075-4

Howden-Chapman, P., Bennett, J., Edwards, R., Jacobs, D., Nathan, K., & Ormandy, D. (2023). Review of the Impact of Housing Quality on Inequalities in Health and Well-Being. Annual Review of Public Health, 44(1), 233–254. 10.1146/annurev-publhealth-071521-111836

Jacobs, D. E., Ahonen, E., Dixon, S. L., Dorevitch, S., Breysse, J., Smith, J., Evens, A., Dobrez, D., Isaacson, M., Murphy, C., Conroy, L., & Levavi, P. (2015). Moving into green healthy housing. J Public Health Manag Pract, 21(4), 345–354. 10.1097/phh.0000000000000047

Jacobs, D. E., Breysse, J., Dixon, S. L., Aceti, S., Kawecki, C., James, M., & Wilson, J. (2014). Health and housing outcomes from green renovation of low-income housing in Washington, DC. J Environ Health, 76(7), 8–16; quiz 60.

Jacobs, D. E., Brown, M. J., Baeder, A., Sucosky, M. S., Margolis, S., Hershovitz, J., Kolb, L., & Morley, R. L. (2010). A systematic review of housing interventions and health: introduction, methods, and summary findings. Journal of Public Health Management and Practice, 16(5), S5–S10.

Jonathan, A. C. S., Miguel, A. H., Barnaby, C. R., Jelena, S., Nancy, D. B., Meera, V., David, H., Douglas, G. A., Mohammed, T. A., Isabelle, B., James, R. C., An-Wen, C., Rachel, C., Jonathan, J. D., Asbjørn, H., Jamie, K., Peter, J., Yoon, K. L., Theresa, D. P.,…Julian, P. T. H. (2016). ROBINS-I: a tool for assessing risk of bias in non-randomised studies of interventions. Bmj, 355, i4919. 10.1136/bmj.i4919

Li, A., Baker, E., & Bentley, R. (2022). Understanding the mental health effects of instability in the private rental sector: A longitudinal analysis of a national cohort. Soc Sci Med, 296, 114778. 10.1016/j.socscimed.2022.114778

Li, A., Baker, E., & Bentley, R. (2025). Housing and mental health inequalities during COVID-19: the role of income and housing support measures. Housing Studies, 40(6), 1379–1400.

Li, A., Toll, M., & Bentley, R. (2023). Health and housing consequences of climate-related disasters: a matched case-control study using population-based longitudinal data in Australia. The Lancet Planetary Health, 7(6), e490–e500. 10.1016/S2542-5196(23)00089-X

Li, A., Toll, M., & Bentley, R. (2024). The risk of energy hardship increases with extreme heat and cold in Australia. Communications Earth & Environment, 5(1), 595.

Liao, Y., Fan, B., Zhang, H., Guo, L., Lee, Y., Wang, W., Li, W., Gong, M., Lui, L., & Li, L. (2021). The impact of COVID-19 on subthreshold depressive symptoms: a longitudinal study. Epidemiology and psychiatric sciences, 30, e20.

Mishra, S. R., Wilson, T., Andrabi, H., Ouakrim, D. A., Li, A., Akpan, E., Bentley, R., & Blakely, T. (2023). The total health gains and cost savings of eradicating cold housing in Australia. Social Science & Medicine, 334, 115954. 10.1016/j.socscimed.2023.115954

Nagare, R., Woo, M., MacNaughton, P., Plitnick, B., Tinianov, B., & Figueiro, M. (2021). Access to Daylight at Home Improves Circadian Alignment, Sleep, and Mental Health in Healthy Adults: A Crossover Study. Int J Environ Res Public Health, 18(19). 10.3390/ijerph18199980

Page, K., Hossain, L., Liu, D., Kim, Y. H., Wilmot, K., Kenny, P., Campbell, M., Cumming, T., Kelly, S., & Longden, T. (2025). Outcomes from the Victorian Healthy Homes Program: a randomised control trial of home energy upgrades. BMJ open, 15(2), e082340.

Poortinga, W., Jones, N., Lannon, S., & Jenkins, H. (2017). Social and health outcomes following upgrades to a national housing standard: a multilevel analysis of a five-wave repeated cross-sectional survey. BMC Public Health, 17(1), 927. 10.1186/s12889-017-4928-x

Sawyer, A., Sherriff, N., Bishop, D., Darking, M., & Huber, J. W. (2022). “It’s changed my life not to have the continual worry of being warm” – health and wellbeing impacts of a local fuel poverty programme: a mixed-methods evaluation. BMC Public Health, 22(1), 786. 10.1186/s12889-022-12994-4

Shishegar, N., & Boubekri, M. (2022). Lighting up living spaces to improve mood and cognitive performance in older adults. Journal of Environmental Psychology, 82, 101845. 10.1016/j.jenvp.2022.101845

Thomson, H., Petticrew, M., & Morrison, D. (2001). Health effects of housing improvement: systematic review of intervention studies. Bmj, 323(7306), 187–190. 10.1136/bmj.323.7306.187

Thomson, H., Thomas, S., Sellstrom, E., & Petticrew, M. (2009). The health impacts of housing improvement: a systematic review of intervention studies from 1887 to 2007. American journal of public health, 99(S3), S681–S692.

Thomson, H., Thomas, S., Sellstrom, E., & Petticrew, M. (2013). Housing improvements for health and associated socioeconomic outcomes. Cochrane Database of Systematic Reviews(2). 10.1002/14651858.CD008657.pub2

Tonn, B., Hawkins, B., Rose, E., Marincic, M., Pigg, S., & Cowan, C. (2023). Saving lives by saving energy? Examining the health benefits of energy efficiency in multifamily buildings in the United States. Building and Environment, 228, 109716. 10.1016/j.buildenv.2022.109716

Tonn, B., Rose, E., Hawkins, B., & Marincic, M. (2021). Health and financial benefits of weatherizing low-income homes in the southeastern United States. Building and Environment, 197, 107847. 10.1016/j.buildenv.2021.107847

Vaid, U. (2023). Physical and mental health impacts of housing improvement: A quasi-experimental evaluation of in-situ slum redevelopment in India. Journal of Environmental Psychology, 86, 101968.

World Health Organization. (2018). WHO housing and health guidelines. World Health Organization. https://apps.who.int/iris/handle/10665/276001

